# Peripheral neuropathy and the limits of personal protective equipment in an agrochemical-exposed mining-farming community in south-east Nigeria: a cross-sectional study

**DOI:** 10.64898/2026.09.21.26363549

**Authors:** Samson I. Abanni

**Author notes:** Correspondence ·.

## Abstract

**Objectives:** To estimate the prevalence of objective peripheral neuropathy, and to test whether self-reported use of personal protective equipment (PPE) is associated with lower neuropathy risk, among adults attending a community hospital in a Nigerian population exposed to both agrochemicals and mining.

**Design:** Clinic-based cross-sectional study (June–November 2025), reported per STROBE.

**Setting:** Infant Jesus Community Hospital, Ishiagu, Ebonyi State, a community combining smallholder farming with granite and lead–zinc mining.

**Participants:** 285 adults (≥18 years) attending the outpatient screening department; people with known diabetes were excluded.

**Main outcome measures:** Peripheral neuropathy on the Michigan Neuropathy Screening Instrument (MNSI) examination — an instrument validated in diabetic populations — defined a priori as an abnormal 10 g monofilament test or absent/reduced ankle reflexes; three alternative case definitions were pre-specified. PPE use was scored 0–12. Associations were estimated by multivariable logistic regression adjusted for age and sex.

**Results:** The proportion screening positive was 47.0% (95% CI 41.2–52.8) under the primary definition and ranged from 29.8% to 56.8% across definitions. PPE use was near-absent (89.1% never used gloves; 43.2% used none) and showed no detectable protective association (adjusted OR 0.97 per point, 95% CI 0.88–1.06). Men used PPE more than twice as often as women (55.1% vs 24.5%, p<0.001), yet neuropathy did not differ by sex (41.6% vs 49.5%, p=0.27). Only age was associated with neuropathy (adjusted OR 1.02 per year, p=0.02).

**Conclusions:** Nearly half of the adults sampled screened positive for peripheral neuropathy, and protective equipment showed no detectable protective association. Because PPE use was near-absent, the study cannot separate ineffective protection from too little protection to measure, and the cross-sectional design precludes causal inference. The equal burden across sexes despite markedly unequal protective behaviour is consistent with community-wide environmental exposure; reducing the total toxic load is a plausible priority that direct exposure measurement should now test.

## Introduction

Pesticide-safety policy across sub-Saharan Africa rests on one rarely examined premise: that the person who handles the chemical is the person at risk, and that shielding that person during application controls the hazard. Acute pesticide poisoning is a major global health burden.[1] The hierarchy of controls ranks personal protective equipment (PPE) last, yet in low-resource farming PPE is promoted as the first, and often the only, measure that reaches smallholders. The premise holds only where exposure is occupational and applicator-bound. Where it is not, the policy guards the wrong thing.

Two features of many rural Nigerian communities strain that premise. First, agrochemicals have shifted from a seasonal input toward a routine tool. In communities such as Ishiagu, herbicides clear land for maize and okra, control weeds around the home, and replace manual labour through the farming year; use is widespread and often without observance of re-entry intervals. Second, farming is not the only neurotoxic source. Ishiagu hosts established granite and lead–zinc mining, layering heavy-metal exposure onto agrochemical exposure in the same soil, water and population. The result resembles a contaminated commons more than a workplace hazard.

Peripheral neuropathy is a sensitive, clinically meaningful endpoint for this cumulative load. Organophosphates injure peripheral nerves through inhibition of neuropathy target esterase and downstream axonal degeneration, and heavy metals such as lead produce a distal, length-dependent sensorimotor neuropathy.[2–5] Self-reported symptoms, which dominate the regional literature, are prone to recall and reporting bias;[6] a structured physical examination detects established nerve injury more directly.[7,8]

Peripheral neuropathy is common in sub-Saharan Africa but is described almost entirely in clinical risk groups. A recent systematic review of 116 studies reported a pooled community prevalence of 4.3%, a pooled hospital/clinic frequency of 39.6%, and about 50% among hospital patients with diabetes.[9] What is missing is an objective estimate from a general adult population in a high-exposure agricultural setting, and a direct test of whether the protective measures that policy promotes actually track with less disease.

We therefore set out to estimate the prevalence of objective peripheral neuropathy among adults attending a community hospital in a mining-farming community, and to test whether higher self-reported PPE use is associated with lower neuropathy risk. We reasoned that the pattern of disease — its size, and its distribution across people who do and do not use protection — is itself informative about where exposure comes from, and therefore about what policy can realistically address. General-population, examination-based neuropathy studies exist in sub-Saharan Africa; what is new here, to our knowledge, is the joint agrochemical-and-mining exposure setting and a direct test of the PPE–neuropathy association in such a population.

## Methods

### Study design and setting

We conducted a clinic-based cross-sectional study between June and November 2025 at Infant Jesus Community Hospital, a non-governmental facility in Ishiagu, Ivo District, Ebonyi State, south-eastern Nigeria. Ishiagu is a dual-economy community in which intensive smallholder farming coexists with established granite and lead–zinc mining, creating overlapping agrochemical and heavy-metal exposure; neither exposure was measured at the individual level in this study. Reporting follows the STROBE statement for cross-sectional studies.[10]

### Participants and sampling

We sampled adults (≥18 years) systematically during outpatient screening sessions, approaching both patients and accompanying relatives so as to capture community members across the spectrum of health rather than an acutely unwell subset, and to limit the healthy-worker and admission-rate biases that affect purely occupational or inpatient cohorts. People with known diabetes mellitus were excluded, because diabetic neuropathy would otherwise dominate the outcome, as were those unable to complete the foot examination. The number screened out for diabetes or an incomplete examination before data entry was not separately logged, which we note as a limitation; the analytic dataset comprised 287 records, from which two participants below 18 years were removed, leaving 285. This is a clinic-based, non-probability sample and not a community probability sample.

We did not perform an a priori sample-size calculation; the sample comprised the eligible attendees recruited over the study window. Because protective-equipment use proved very low (see Results), statistical power to detect a protective effect of PPE was limited — a point we return to in the discussion.

### Outcome assessment

A single trained clinician performed the Michigan Neuropathy Screening Instrument physical examination on every participant, ensuring consistent technique. We defined peripheral neuropathy a priori as an abnormality on the 10 g monofilament test at two or more sites (loss of protective sensation) or absent/reduced ankle reflexes. This definition omits vibration perception, which declines with age independently of toxic injury, to favour signs more specific to clinically important damage. Because any single threshold is a judgement, we pre-specified three alternative definitions for sensitivity analysis: any one of the three signs abnormal (most sensitive); two or more signs abnormal (most specific); and loss of protective sensation alone.[7]

Two cautions follow from the instrument. The MNSI was developed and validated in diabetic populations;[7,8] its operating characteristics in a non-diabetic, mixed-aetiology, agrochemical- and metal-exposed population are not established, so our estimates represent the proportion screening positive on this tool rather than a validated neuropathy prevalence. The single examiner was not formally blinded to participants’ self-reported PPE use, recorded at the same encounter, which is a potential source of observer bias in the central exposure comparison.

### Exposure assessment

Participants reported how often they used each of four protective items — gloves, mask or respirator, boots and long protective clothing — on a four-level scale (never, rarely, sometimes, always) scored 0–3 and summed to a composite of 0–12. We analysed the composite continuously, in ordered categories (none, 0; minimal, 1–3; moderate, 4–6; good, 7–12) and as a high-versus-low contrast; glove use was examined separately as the barrier most relevant to dermal absorption.

The instrument did not record chemical type, application frequency, mixing practices, water source or mining proximity, so community-level agrochemical exposure and mining are described as context, not measured at the individual level.

### Statistical analysis

Continuous variables are summarised as mean ± standard deviation or median (interquartile range), and categorical variables as counts and percentages. Prevalence is reported with a 95% confidence interval (Wald) under each case definition. Bivariate associations used the chi-square test (with Yates correction for 2×2 tables) and the Cochran–Armitage test for trend across age groups. A multivariable logistic regression model for the primary outcome included age (centred), sex and the PPE composite, with adjusted odds ratios (aOR) and 95% confidence intervals. Age was kept continuous after the linearity-of-logit assumption was met. The primary outcome, the three alternative case definitions, and the age+sex+PPE model were pre-specified; analyses beyond that model (high glove use, the age-by-sex interaction, the stringent-definition refit) are exploratory, and we did not adjust for multiplicity. Analysis used Python (NumPy) with maximum-likelihood logistic regression; two-sided p<0.05 was treated as significant.

### Ethics

The study followed the Declaration of Helsinki and was approved by the Ebonyi State Ministry of Health Research Ethics Committee (approval number: [INSERT APPROVAL NUMBER]), with institutional permission from Infant Jesus Community Hospital. The committee approved verbal (rather than written) informed consent, which was appropriate to a non-invasive questionnaire- and-examination design and to the low literacy levels in the population. No personal identifiers (names, addresses or telephone numbers) were collected, so no participant can be identified from the data.

## Results

### Participant characteristics

Of 285 adults, 196 (68.8%) were women and the mean age was 44.3 ± 11.9 years (range 18–82); the population was predominantly Igbo. Educational attainment ranged from none (16.1%) to tertiary (24.2%), with one record (0.4%) unclassifiable. Protective-equipment use was low throughout: the mean composite score was 2.5 of a possible 12, 43.2% used no protection at all, and only 9.8% reached the good-use category. Glove use, the barrier most relevant to dermal absorption, was almost absent — 89.1% never used gloves (Table 1).

**Table 1.**
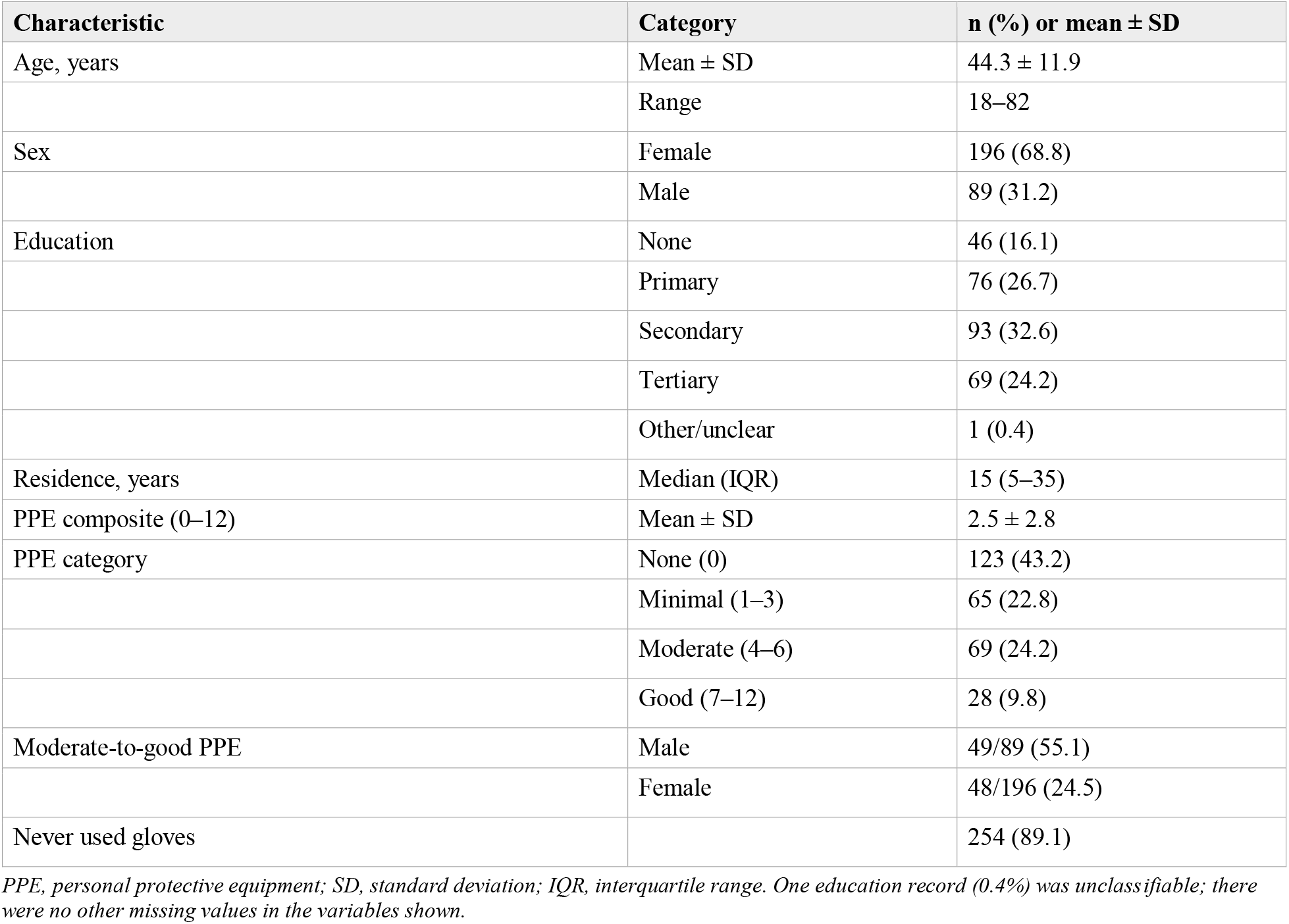
Characteristics of participants (N = 285).

### Prevalence of peripheral neuropathy

Under the primary definition, 134 of 285 participants screened positive for peripheral neuropathy: 47.0% (95% CI 41.2–52.8). Estimates ranged from 29.8% for loss of protective sensation alone to 56.8% when any single sign was counted (Table 2). Impaired vibration perception (45.3%) and reduced ankle reflexes (38.2%) were the commonest findings. Almost a third of participants (30.5%) had a visible foot deformity, dry skin or ulcer on inspection — a non-specific sign with many causes, reported here for completeness and not counted in any neuropathy definition. In a non-diabetic general adult sample, these figures fall within the clinic-based range reported for sub-Saharan Africa.[9]

**Table 2.**
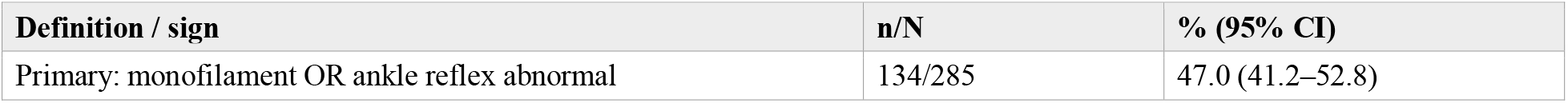
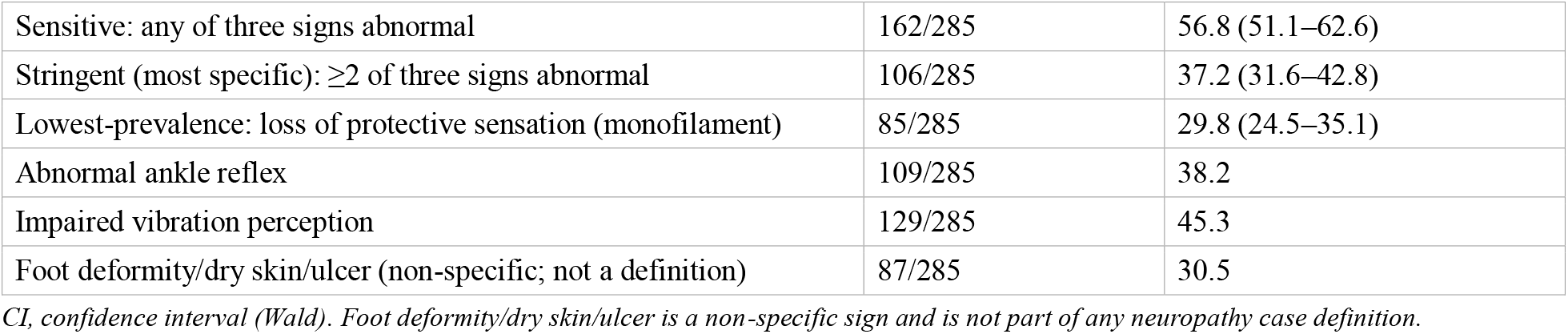
Proportion screening positive for peripheral neuropathy by case definition and examination sign (N = 285).

### Age, but not protection, tracks with disease

Prevalence rose across the working-age span, from 31.0% in those under 30 to 60.0% at 40–49, before easing in the oldest group, giving a significant trend (Cochran–Armitage z=2.37, p=0.018; Table 3). The fall after 60 may reflect a survivor effect or the smaller number at that age and should not be over-read. PPE category showed no graded relationship with neuropathy: prevalence was 50.4% in those using no PPE and 35.7% in the good-use group, with no significant difference across categories (Pearson χ^2^=3.17, df=3, p=0.37) and no monotonic trend.

**Table 3.**
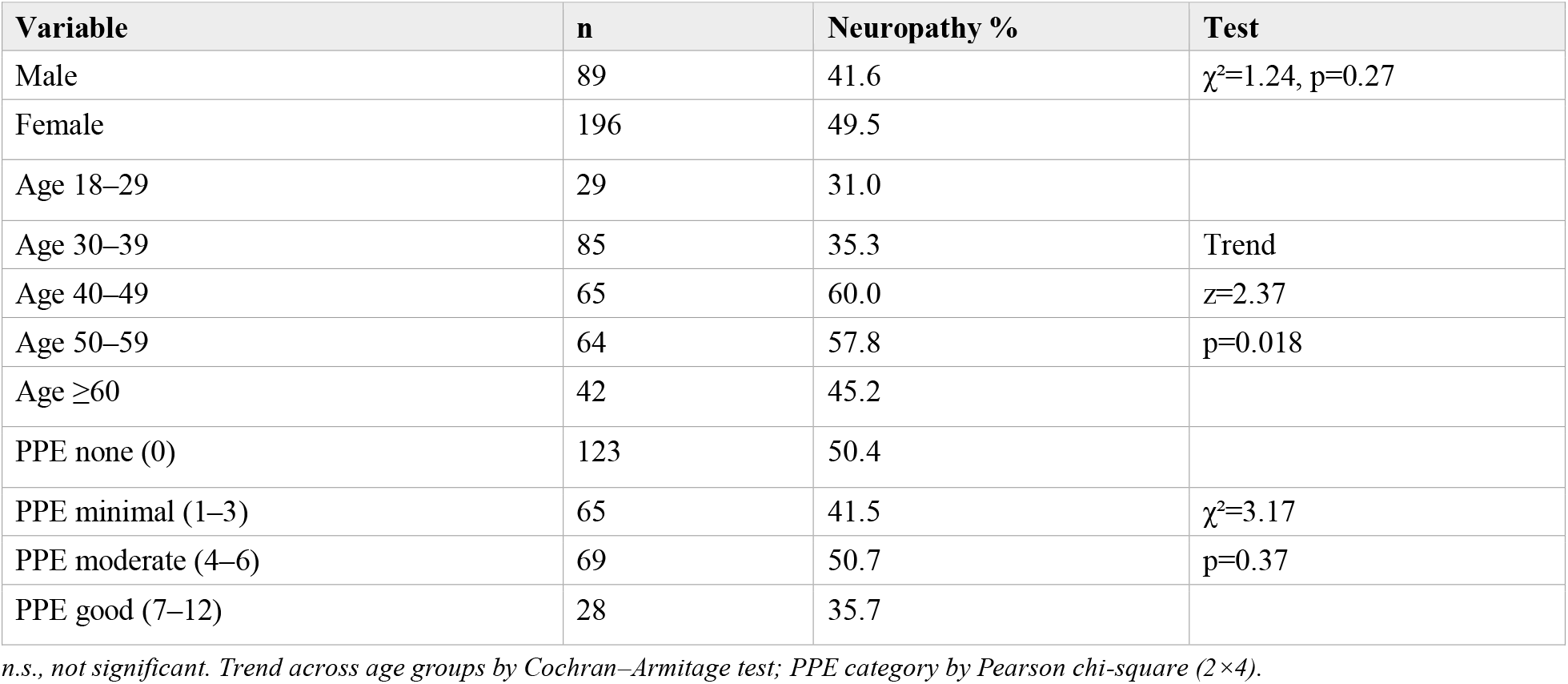
Neuropathy by sex, age group and PPE category (primary definition).

### Unequal protection, equal disease

Protective behaviour differed sharply by sex. Men used moderate-to-good PPE more than twice as often as women (49/89, 55.1% vs 48/196, 24.5%; χ^2^=24.1, p<0.001), in keeping with a division of labour in which men spray with boots and masks while women weed and harvest with little protection. Disease did not follow behaviour. Neuropathy was, if anything, commoner in women (49.5%) than men (41.6%), and the difference was not significant (p=0.27). The group that protected itself least was no worse affected than the group that protected itself most.

### Multivariable analysis

After adjustment for age and sex, the PPE composite was not associated with neuropathy (aOR 0.97 per point, 95% CI 0.88–1.06, p=0.50), and isolating high glove use gave the same null result (aOR 0.76, 95% CI 0.35–1.68, p=0.50; Table 4). The confidence intervals are wide and admit both modest protection and modest harm; given how few participants used substantial protection (mean 2.5/12; 89% never used gloves), the study had limited power to detect protection even had it existed. Age was the only predictor associated with the outcome (aOR 1.02 per year, 95% CI 1.00– 1.05, p=0.02). The results held when the outcome was redefined as two or more abnormal signs (PPE aOR 0.97, p=0.58; age aOR 1.03, p=0.01); a pre-specified age-by-sex interaction was not significant (likelihood-ratio p=0.63); and model discrimination was modest (area under the curve 0.59), meaning individual-level prediction is weak — as expected when the dominant exposure is shared across the population and carries little between-person signal. These secondary analyses are exploratory.

**Table 4.** Logistic regression for peripheral neuropathy (primary definition; N = 285).

| Predictor | Crude OR (95% CI) | Adjusted OR (95% CI) | p |
| --- | --- | --- | --- |
| Age (per year) | 1.03 (1.01–1.05) | 1.02 (1.00–1.05) | 0.02 |
| Female sex | 1.38 (0.83–2.28) | 1.24 (0.72–2.14) | 0.43 |
| PPE score (per point) | 0.94 (0.87–1.03) | 0.97 (0.88–1.06) | 0.50 |
| High glove use* | — | 0.76 (0.35–1.68) | 0.50 |

OR, odds ratio; CI, confidence interval. \*Modelled separately, replacing the PPE composite, adjusted for age and sex. High glove use and the interaction term are exploratory.

## Discussion

Nearly half of adults attending this mining-farming community hospital screened positive for peripheral neuropathy, in a sample from which diabetes had been excluded. Protective equipment was scarce and, where used, showed no measurable association with lower disease. Women — who used far less PPE and rarely sprayed — carried a burden equal to or greater than that of men. Age was the only individual-level correlate. Read together, these findings sit awkwardly with the occupational model, in which risk should concentrate where the protective policy is aimed.

The distribution of disease is the argument, and it is an indirect one. If neurotoxic exposure occurred mainly during application, those who spray most and protect least should bear the heaviest burden, and PPE should bend the curve. Neither held. Equal burden across sexes, despite a two-fold difference in protective behaviour, is difficult to reconcile with applicator-bound exposure and is consistent with exposure that reaches the whole community — through herbicide-laden soil worked during weeding, contaminated water, drift into homes, premature re-entry, and residues carried home on clothing and equipment. Onto this is layered the heavy-metal burden of granite and lead–zinc mining, which shares the same soil and water and produces a clinically indistinguishable distal neuropathy.[5] In such a setting the unit of exposure may be the environment rather than the individual, and a barrier worn only while spraying would address a fraction of the dose. This remains a hypothesis: rival explanations — sex differences in other neuropathy risks, the non-specific outcome, and residual confounding — are not excluded by these data.

The null PPE result must be read with its variance, not as proof of futility. A protective device can fail to register either because it does not work against the operative route or because too few people use it to reveal an effect; here both may apply. Protection was almost absent where it matters most — 89% never used gloves, leaving the hands, the main route of dermal absorption, unguarded even when masks or boots were worn — so the study is structurally underpowered to detect protection regardless of whether PPE works. Where equipment is used it is typically dust-grade rather than chemical-resistant, is reused beyond its useful life, and under tropical heat becomes soaked with sweat and pesticide, so that contaminated gear may hold toxin against the skin rather than exclude it.[11] And in a cross-sectional design any weak protective signal is liable to reverse causation, as those already affected take up protection only after symptoms begin.

A prevalence of 47% needs a careful benchmark. Community-based neuropathy prevalence in sub-Saharan Africa pools at roughly 4%, but clinic-based frequencies run near 40% and reach about 50% among hospital patients with diabetes.[9] Our estimate is a clinic-based, screening-level figure and should be read as such; the examination is a screen calibrated in diabetic cohorts and under-detects relative to nerve conduction studies rather than over-calling.[7,8] The notable feature is not that the figure towers over a general-population baseline, but that a non-diabetic agricultural population reaches the frequency usually seen in diabetic clinic cohorts. The range we report — 29.8% under the lowest-prevalence definition (loss of protective sensation) and 37.2% under the most specific (two or more signs) — brackets the uncertainty honestly and still describes a heavy, under-recognised burden. The scarcity of protective equipment matches other Nigerian and West African surveys, in which most farmers never use respirators, gloves or boots.[12]

The implication, if exposure is indeed environmental, is structural. No achievable level of individual compliance can resolve a problem of this size, and compliance campaigns risk shifting responsibility onto the people least able to control the hazard. The hierarchy of controls places elimination, substitution and engineering controls above PPE; policy here has inverted that order. Our data are consistent with prioritising reduction of the total toxic load — restriction of the most hazardous (WHO Class I) agrochemicals, enforcement of re-entry intervals and control of mining effluent — alongside protection of water sources and population-level biomonitoring.[13] PPE retains a role for the applicator while mixing and spraying; the case made here is that it cannot be assumed sufficient as the centrepiece of safety policy for a contaminated commons, and that direct exposure measurement should test this.

### Strengths and limitations

The main strength is an objective, examiner-based outcome applied by one clinician to a broad sample that includes the relatively well, reducing reliance on symptom self-report and limiting healthy-worker and admission-rate biases. Pre-specified alternative case definitions and exposure measures show the conclusions are not artefacts of a single analytic choice.

The limitations are substantial and bound every inference above. First, a floor effect: with mean PPE 2.5/12 and 89% never using gloves, the study is underpowered to detect protection even if PPE were effective, so the null cannot distinguish “PPE does not help” from “too little PPE use to show that it helps.” Second, the regression adjusts only for age and sex; major neuropathy risk factors — alcohol, vitamin B12 status, HIV, leprosy, height, farming intensity, socioeconomic position and the mining exposure itself — were unmeasured. This both weakens the prevalence interpretation and means PPE users may differ systematically (wealthier, more educated, less manual field labour), which could mask real protection; the environmental-commons reading rests partly on the absence of these measurements and is weakened accordingly. Third, the MNSI is validated for diabetic neuropathy, so our figure is a screen-positive proportion, not a validated prevalence, and the single examiner was not formally blinded to PPE status. Fourth, exposure to agrochemicals and to mining was not quantified — no chemical type, application intensity, proximity, water source, or biomarkers such as blood cholinesterase or blood lead — so the environmental interpretation is an inference from the pattern of disease, not a measurement. Fifth, this is a clinic-based, non-probability sample with women over-represented (69%), and the recruited-to-analysed cascade was incompletely logged. Sixth, multiple case definitions and PPE parameterisations were examined; pre-specification mitigates but does not remove the resulting multiplicity, and the non-primary analyses are exploratory. We could not exclude undiagnosed diabetes, B12 deficiency, HIV, alcohol use or leprosy; these are unlikely to vary systematically with PPE use, however, and so are an improbable explanation for the specific null association. Finally, the examination is a screen, not a confirmatory nerve conduction study.

## Conclusion

In a community where agrochemicals are used widely and mining adds a second neurotoxic source, close to half of the adults sampled screened positive for peripheral neuropathy, and the protective equipment that policy promotes neither reached most people nor, where used, was associated with lower risk. Equal disease across sexes despite unequal protection is consistent with exposure that is shared across the community rather than confined to the sprayer, though this study cannot prove it: PPE use was too low to test protection fairly, and exposure was not measured directly. The defensible next step is a community-based study with environmental sampling of soil and water, individual biomarkers of agrochemical and heavy-metal exposure, and confirmatory electrophysiology, ideally followed over time — work that would establish whether reducing the total toxic load, rather than individual protection, is what this population needs.

## Declarations

## Ethics approval and consent

The study followed the Declaration of Helsinki and was approved by the Ebonyi State Ministry of Health Research Ethics Committee (approval number: [INSERT APPROVAL NUMBER]), with institutional permission from Infant Jesus Community Hospital. The committee approved verbal (rather than written) informed consent, appropriate to a non-invasive questionnaire-and-examination design and to low literacy in the population. No personal identifiers were collected, so no participant can be identified from the data.

## Funding

Self-funded by the authors; no external or commercial sponsorship.

## Competing interests

None declared.

## Data availability

The de-identified dataset and analysis code are available from the corresponding author on reasonable request.

## Use of artificial intelligence

The authors used a large language model (Claude, Anthropic) to assist with drafting, editing and statistical-analysis scripting. The authors designed the study, verified all analyses and outputs, and take full responsibility for the content. No AI tool is listed as an author.

## Author contributions

SIA conceived and designed the study, supervised data collection, performed the clinical examinations, analysed the data and drafted the manuscript.

## Notes

### Competing Interest Statement

The authors have declared no competing interest.

### Author Declarations

The study followed the Declaration of Helsinki and was approved by the Ebonyi State Ministry of Health Research Ethics Committee, with institutional permission from Infant Jesus Community Hospital.

